# Scalable biological-cognitive profiling for Alzheimer’s disease in the population

**DOI:** 10.1101/2025.08.20.25333966

**Authors:** Karin Lohi, Aino Aaltonen, Sanna-Kaisa Herukka, Tarja Kokkola, Sari Kärkkäinen, Mia Urjansson, FinnGen, Aarno Palotie, Heiko Runz, Jaakko Kaprio, Valtteri Julkunen, Toni Saari, Eero Vuoksimaa

## Abstract

Plasma phosphorylated tau217 has been suggested as a core biomarker for establishing biological Alzheimer’s disease diagnosis. This blood biomarker has not been studied together with scalable cognitive assessment tools in population-based samples. We investigated the prevalence of cognitive and Alzheimer’s disease biomarker abnormality and associations between plasma phosphorylated tau217 and remotely measured cognitive function in individuals without dementia.

We used a population-based cross-sectional sample of 65–85-year-olds (n=692, 57% females) excluding those with previously diagnosed Alzheimer’s disease or other dementia-causing neurodegenerative disease. Cognition was measured with telephone-administered word list recall task (episodic memory) and animal naming (semantic fluency). Plasma phosphorylated tau217 was determined with the ALZpath assay.

The prevalence of individuals with abnormalities in tests measuring episodic memory, semantic fluency and plasma phosphorylated tau217 was 10–13%. Higher plasma phosphorylated tau217 was related to lower scores in telephone-administered cognitive tests.

We found a substantial minority of a population-based sample of individuals without a clinical diagnosis of Alzheimer’s disease to have cognitive and plasma phosphorylated tau217 profiles suggesting underlying Alzheimer’s disease. Combining plasma phosphorylated tau217 with remote cognitive assessment could be a scalable, accessible, and cost-effective protocol for screening individuals with undiagnosed or risk for Alzheimer’s disease.

## Introduction

The current clinical practice often results in late detection of Alzheimer’s disease (AD) with a substantial proportion of patients diagnosed at moderate or severe stage.^1^ Early diagnosis would benefit patients because current treatments are efficient in early stages of the disease^2^ and reduce healthcare costs considerably through better disease management.^3^ Moreover, drug and intervention trials would benefit from improved screening of early AD. Developing a scalable cost-effective approach for early detection, diagnosis and monitoring of AD is needed.

Plasma phosphorylated tau217 (p-tau217) – reflecting both amyloid beta (Aβ) and tau pathologies indicating Alzheimer’s disease neuropathological changes (ADNPC) – has been suggested as an accurate and early core biomarker sufficient to diagnosis of AD independently of clinical symptoms.^4–7^ However, it has also been argued that AD diagnosis should not be based solely on AD biomarkers and that biomarker status should not be determined in cognitively healthy individuals as many of them are unlikely to develop symptoms in the next couple of years.^8^ In reality, many individuals who are considered “cognitively healthy” may have mild or even substantial cognitive impairment simply because their cognition has not been assessed.

The major advantages of plasma biomarkers over cerebrospinal fluid samples and PET-imaging are accessibility, cost-effectiveness, and applicability for large-scale use, but research in population-based samples of cognitively normal individuals remains limited.^9^ Remote cognitive assessment with unsupervised digital memory assessment together with p-tau217 has been reported to have prognostic value^10^, but not all digital tools are scalable at population level because they require access and knowledge of appropriate software. Telephone interview would be more practical alternative for remote cognitive testing, also in low- and middle-income countries. Episodic memory and semantic fluency, cognitive domains that best reflect early AD-related cognitive decline and predict progression in the AD continuum^11^ can be assessed remotely via telephone interview^12,13^ but to our knowledge they have not been used together with plasma p-tau217 previously.

By combining cognitive telephone interview and blood sampling, the aim of this study was to examine the prevalence of abnormal p-tau217 and cognitive impairment using low-cost, scalable methods in a population-based sample in individuals without a clinical diagnosis of AD or other neurodegenerative disease. We also examined the continuous associations between telephone-administered cognitive tests and plasma p-tau217.

## Methods

### Participants

Participants were 65–85-year-olds from a cross-sectional population-based biobank recall study called TWINGEN with data collection in 2023.^14^ TWINGEN excluded individuals with AD, other neurodegenerative disease or other cognition-affecting disease based on health registry data diagnosis.^14^ We studied 692 individuals who gave blood samples and participated in the telephone interview assessing cognitive performance (Supplementary Figure 1). TWINGEN study has ethical approval from the Helsinki and Uusimaa hospital district (HUS) Regional Committee on Medical Research Ethics (approval number 16831/2022). In addition, permission to recontact sample donors through the biobank was granted from THL Biobank (diary ID 83/2022). All participants provided written informed consent.

### Plasma p-tau217

Plasma was extracted from a non-fasting blood samples and, aliquots were sent to the Biomarker Laboratory of the University of Eastern Finland for quantifying p-tau217 with ALZpath Simoa pTau-217 v2 Assay Kit (Quanterix, Ref# 104371).^15^ We used a Box-Cox - transformed^16^ continuous plasma p-tau217 measure and categories based on two validated cut-offs: (>0.42 pg/mL)^15^ and (>0.475 pg/mL)^17^ to indicate dichotomic Aβ positivity (ADNPC that implies biologically defined AD). Additionally we used a three-category ADNPC classification of plasma p-tau217 levels —low, intermediate, and high— based on cut-offs from Ashton et al.^15^ (<0.40, 0.40–0.63, >0.63 pg/mL) and Figdore et al.^17^ (<0.405, 0.405–0.590, >0.590 pg/mL).

### Cognitive measures

We used a telephone-administered interview to assess cognition. Episodic memory was measured with validated three trial 10-word list learning task^18^ using immediate (total number of words in trials 1-3, score 0—30) and delayed recall (number of words in free delayed recall, score 0—10) measures^18^. Semantic fluency was measured with one minute animal naming (number of animals).^18^

We used cognitive cut-offs from corresponding in-person measures from the Finnish education-adjusted established norms for the Consortium to Establish a Registry for Alzheimer’s Disease – neuropsychological battery (CERAD-nb) to identify cognitive imapairment^19^ and means (immediate recall = 14.5, delayed recall = 3.8, and semantic fluency = 15.1) from an independent sample of individuals with AD^18^ as reference to compare participant performance to typical AD performance. In addition to categorical cognitive status, we investigated the p-tau217-cognition associations based on continuous measures. Finally, we selected delayed recall and semantic fluency to classify individuals based on the number of tests with impaired scores, i.e., impairment in zero, one or two of these measures based on their significant associations to p-tau in prior analyses.

### Statistical analyses

The descriptive statistics and sex differences were reported with Chi-squared and two-tailed T-tests. The prevalence of cognitive impairment was assessed using Chi-squared tests across dichotomic (normal/abnormal) and three-category (low/intermediate/high) p-tau217 groups. Differences in continuous plasma p-tau217 were examined with Wald-test. Benjamini-Hochberg procedure was applied to all total three-category p-tau217 score and prevalence post-hoc tests.

Associations between p-tau217 and cognitive tests were analysed through Pearson correlations. Linear regression was then used to examine plasma p-tau217, age (centred around the sample mean) and plasma p-tau 217 by age interactions as predictors of cognitive measures.

Statistical significance was considered as p < 0.05. All analyses accounted for participants relatedness (twins within families) by using with *survey* R package.^20^ We followed the Strengthening the Reporting of Observational Studies in Epidemiology guidelines for cross-sectional study.

## Results

### Sample characteristics

The mean age of the participants was 76.17 (SD = 4.57) years, they had median education of 10 years (IQR = 6) and 29% were *APOE* ε4-carriers with no sex differences (Table 1). Female participants performed better in immediate (p < 0.001) and delayed recall (p < 0.001; Table 1). The prevalence of Aβ positivity was 30 % by Figdore et al.^17^ and 38 % by Ashton et al.^15^ cut-offs and did not differ by sex. According to the three-category approach, most participants had low plasma p-tau217 whereas approximately one fifth had high plasma p-tau217 and high probability of ADNPC (Table 1).

**Table 1:**
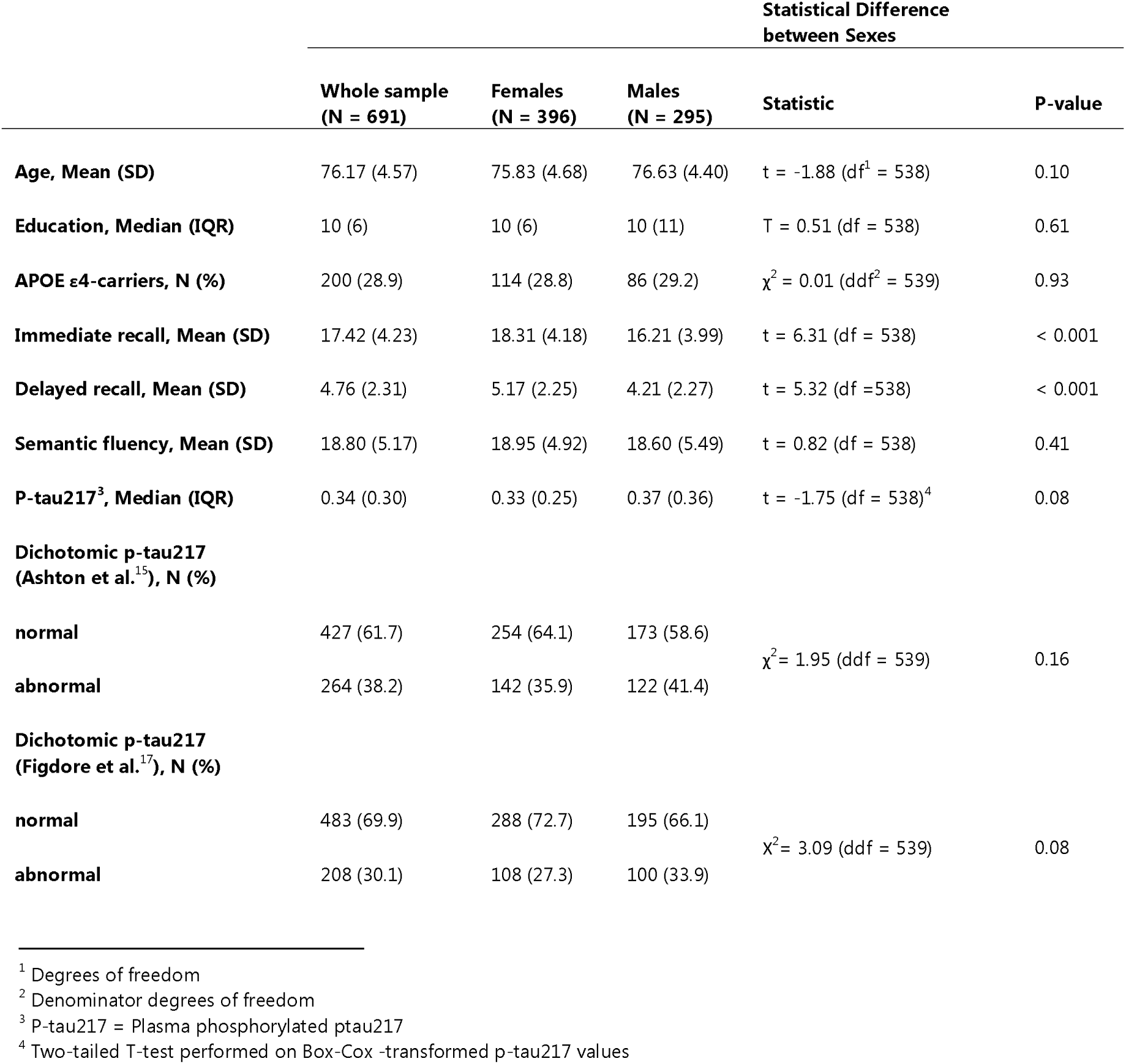
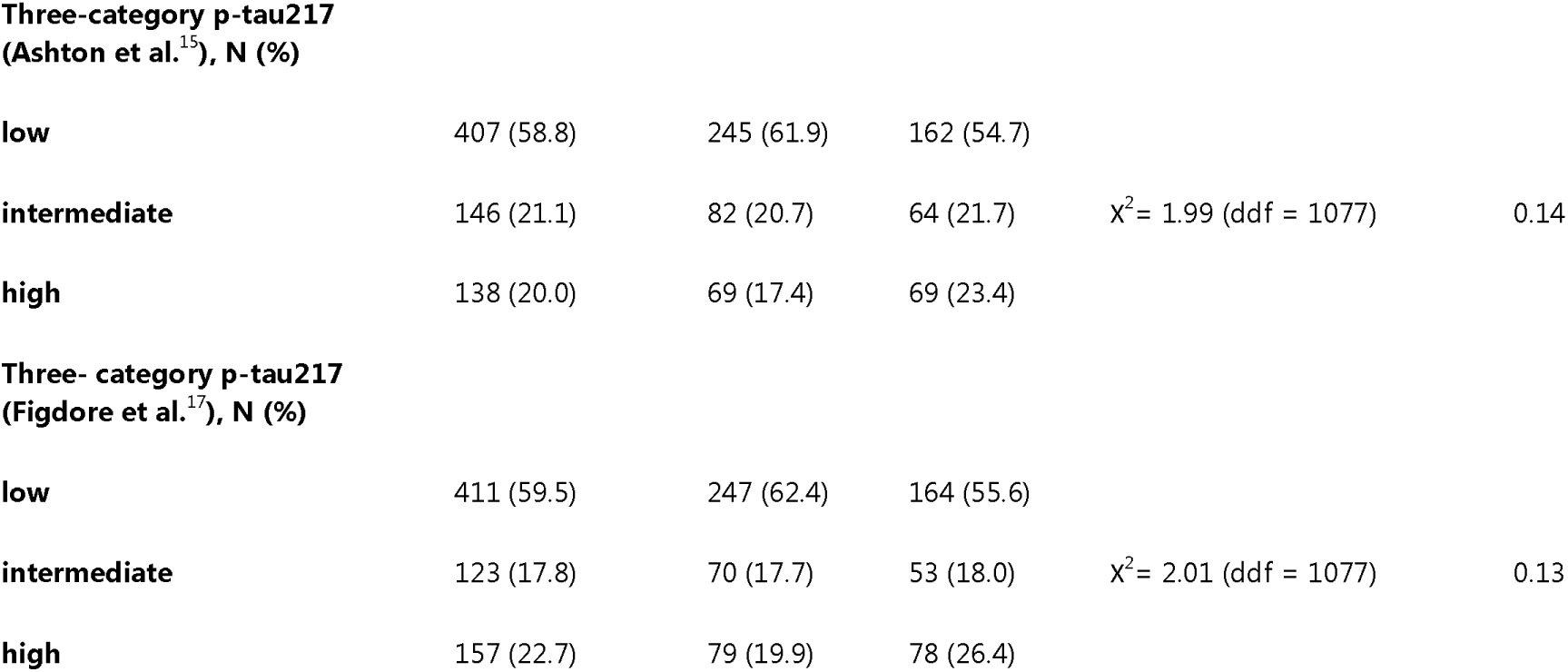
Sample characteristics in the whole sample and by sex.

|  | Statistical Difference<br>between Sexes |  |  |  |  |
| --- | --- | --- | --- | --- | --- |
|  | Whole sample<br>(N = 691) | Females<br>(N = 396) | Males<br>(N = 295) | Statistic | P-value |
| Age, Mean (SD) | 76.17 (4.57) | 75.83 (4.68) | 76.63 (4.40) | t = -1.88 (df<sup>1</sup> = 538) | 0.10 |
| Education, Median (IQR) | 10 (6) | 10 (6) | 10 (11) | T = 0.51 (df = 538) | 0.61 |
| APOE ε4-carriers, N (%) | 200 (28.9) | 114 (28.8) | 86 (29.2) | χ<sup>2</sup> = 0.01 (ddf<sup>2</sup> = 539) | 0.93 |
| Immediate recall, Mean (SD) | 17.42 (4.23) | 18.31 (4.18) | 16.21 (3.99) | t = 6.31 (df = 538) | < 0.001 |
| Delayed recall, Mean (SD) | 4.76 (2.31) | 5.17 (2.25) | 4.21 (2.27) | t = 5.32 (df = 538) | < 0.001 |
| Semantic fluency, Mean (SD) | 18.80 (5.17) | 18.95 (4.92) | 18.60 (5.49) | t = 0.82 (df = 538) | 0.41 |
| P-tau217<sup>3</sup>, Median (IQR) | 0.34 (0.30) | 0.33 (0.25) | 0.37 (0.36) | t = -1.75 (df = 538)<sup>4</sup> | 0.08 |
| Dichotomic p-tau217<br>(Ashton et al.<sup>15</sup>), N (%) |  |  |  |  |  |
| normal | 427 (61.7) | 254 (64.1) | 173 (58.6) | χ<sup>2</sup> = 1.95 (ddf = 539) | 0.16 |
| abnormal | 264 (38.2) | 142 (35.9) | 122 (41.4) |  |  |
| Dichotomic p-tau217<br>(Figdore et al.<sup>17</sup>), N (%) |  |  |  |  |  |
| normal | 483 (69.9) | 288 (72.7) | 195 (66.1) | χ<sup>2</sup> = 3.09 (ddf = 539) | 0.08 |
| abnormal | 208 (30.1) | 108 (27.3) | 100 (33.9) |  |  |

<sup>1</sup> Degrees of freedom
<sup>2</sup> Denominator degrees of freedom
<sup>3</sup> P-tau217 = Plasma phosphorylated ptau217
<sup>4</sup> Two-tailed T-test performed on Box-Cox -transformed p-tau217 values

**Three-category p-tau217 (Ashton et al.<sup>15</sup>), N (%)**
|  |  |  |  |  |  |
| --- | --- | --- | --- | --- | --- |
| **low** | 407 (58.8) | 245 (61.9) | 162 (54.7) |  |  |
| **intermediate** | 146 (21.1) | 82 (20.7) | 64 (21.7) | $\chi^2 = 1.99$ (ddf = 1077) | 0.14 |
| **high** | 138 (20.0) | 69 (17.4) | 69 (23.4) |  |  |
| **Three- category p-tau217 (Figdore et al.<sup>17</sup>), N (%)** |  |  |  |  |  |
| **low** | 411 (59.5) | 247 (62.4) | 164 (55.6) |  |  |
| **intermediate** | 123 (17.8) | 70 (17.7) | 53 (18.0) | $\chi^2 = 2.01$ (ddf = 1077) | 0.13 |
| **high** | 157 (22.7) | 79 (19.9) | 78 (26.4) |  |  |

### Prevalence of concurrent ADNCP and cognitive impairment

Cognitive scores were significantly different between low and high ADNPC groups for delayed recall (t = −3.00, p = 0.003) and semantic fluency (t = −4.08, p < 0.001), but not for immediate recall (t = −2.00, p = 0.05). Impairment in all three cognitive measures was significantly more common in those with ADNPC and performance above mean AD score was associated with normal plasma p-tau217 (Table 2). Results reproduced with Figdore et al. cut-offs (Supplementary Table 1) were similar to those obtained with Ashton et al. cut-offs (Table 2).

**Table 2:** Cognitive performance variance by Ashton et al.^14^ p-tau217 categories.

|  |  | Dichotomic Plasma P-tau217 |  |  | Three-category Plasma P-tau217 |  |  |  |
| --- | --- | --- | --- | --- | --- | --- | --- | --- |
|  |  | Normal | Abnormal | Statistical Difference<br>(statistic [df/ddf], p-value) | Low | Intermediate | High | Statistical Difference<br>(statistic [ddf], p-value) |
| **N** |  | 427 | 264 |  | 407 | 146 | 138 |  |
| **P-tau217 pg/ml, M (SD)** |  | 0.27 (0.08) | 0.73 (0.33) |  | 0.26 (0.08) | 0.49 (0.07) | 0.94 (0.34) |  |
| **Immediate Recall** | **Score, M (SD)** | 17.68 (4.17) | 16.98 (4.29) | t = 1.99 (538), p = 0.05 | 17.72 (4.21) | 17.07 (4.30) | 16.88 (4.16) | W = 2.39 (ddf = 537), p = 0.09 |
| | **Impaired Cognition, N (%)** | 183 (42.9) | 141 (53.2) | $\chi^2 = 6.25$ (539), p = 0.01 | 172 (42.3) | 75 (51.4) | 77 (55.4) | $\chi^2 = 3.96$ (ddf = 1077), p = 0.02 |
| | **Score > AD score, N (%)** | 322 (75.4) | 179 (67.5) | $\chi^2 = 5.09$ (539), p = 0.02 | 308 (75.7) | 102 (70.0) | 91 (65.5) | $\chi^2 = 3.00$ (ddf = 1078), p = 0.05 |
| **Delayed Recall** | **Score, M (SD)** | 4.98 (2.21) | 4.40 (2.42) | t = 3.00 (538), p = 0.003 | 5.00 (2.21) | 4.54 (2.25) | 4.29 (2.56) | W = 4.72 (ddf = 537), p = 0.009 |
| | **Impaired Cognition, N (%)** | 210 (49.2) | 158 (59.6) | $\chi^2 = 6.58$ (539), p = 0.01 | 200 (49.1) | 83 (56.8) | 85 (61.2) | $\chi^2 = 3.26$ (ddf = 1073), p = 0.04 |
| | **Score > AD score, N (%)** | 312 (73.1) | 174 (65.7) | $\chi^2 = 4.30$ (539), p = 0.04 | 299 (73.5) | 99 (67.8) | 88 (63.3) | $\chi^2 = 2.87$ (ddf = 1077), p = 0.06 |
| **Semantic Fluency** | **Score, M (SD)** | 19.44 (5.19) | 17.75 (4.98) | t = 4.01 (538), p < 0.001 | 19.52 (5.13) | 17.90 (5.58) | 17.58 (4.47) | W = 9.82 (ddf = 537), p < 0.001 |
| | **Impaired Cognition, N (%)** | 172 (40.3) | 133 (50.2) | $\chi^2 = 5.86$ (539), p = 0.02 | 161 (39.6) | 78 (53.4) | 66 (47.5) | $\chi^2 = 4.27$ (ddf = 1077), p = 0.01 |
| | **Score > AD score, N (%)** | 326 (76.3) | 176 (66.4) | $\chi^2 = 5.83$ (539), p = 0.02 | 314 (77.1) | 92 (63.0) | 96 (69.1) | $\chi^2 = 2.89$ (ddf = 1076), p = 0.06 |

Delayed recall (W = 4.72, p = 0.009) and semantic fluency (W = 9.82, p < 0.001) scores differed significantly across the three-category ADNPC groups (Table 2). Post-hoc tests yielded significant differences in delayed recall (low-intermediate: t = −2.07, p = 0.12, low-high: t = −2.73, p = 0.02, intermediate-high: t = −0.86, p = 0.49) and in semantic fluency (low-intermediate: t = −3.03, p = 0.01, low-high: t = −3.98, p < 0.001, intermediate-high: t = −0.44, p = 0.71) across three-category ADNPC in. No significant group differences were evident in immediate recall (W = 2.39, p = 0.09; Supplementary Table 2). Cognitive impairment with all three cognitive measures was associated with higher p-tau217 levels based on three-category classification (p’s <0.05; Table 2). Performance above the mean AD score was not associated with three-category ADNPC (immediate: Χ^2^ = 3.00, p = 0.05, delayed: Χ^2^ = 2.87, p = 0.06, semantic fluency: Χ^2^ = 2.89, p = 0.06; Table 2).

### Continuous associations of plasma p-tau217 with cognition

Higher p-tau217 was associated with significantly poorer performance in delayed recall (r = - 0.113, p = 0.006) and semantic fluency (r = −0.167, p < 0.001) but not in immediate recall (r = −0.083, p = 0.052; Supplementary Table 3). In linear regression analyses, p-tau217 was not a significant predictor of immediate recall or delayed recall when adjusting for age (Figure 1). Plasma p-tau217 was negatively associated with semantic fluency in the age-adjusted model (β = –1.251, 95%CI [–1.916, –0.587, p < 0.001; Figure 1). There were no significant p-tau217 by age interactions on any of the cognitive measures (Supplementary Tables 4-5).

**Figure 1:**
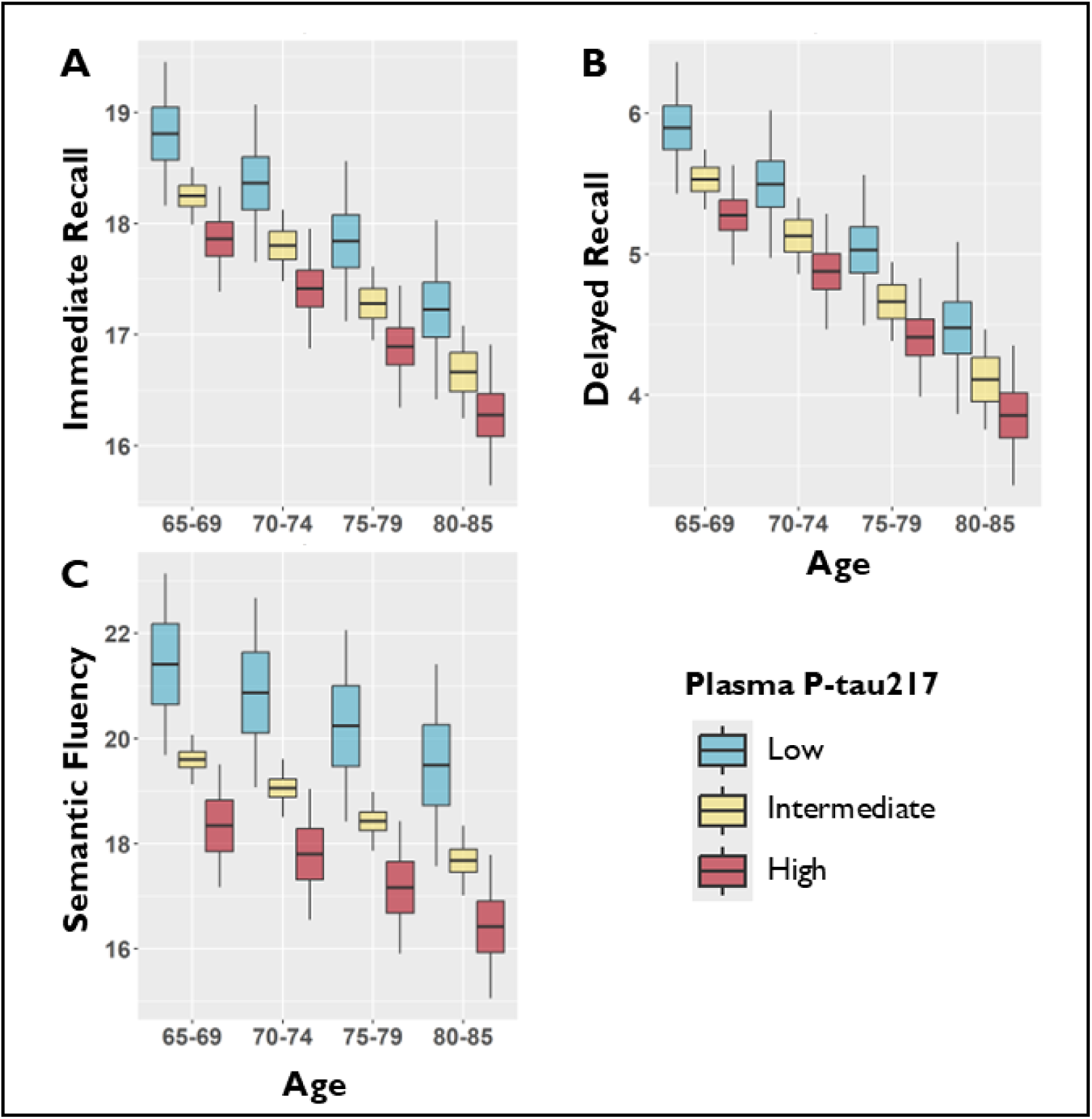
Linear regression predicted cognitive measures by age and p-tau217. Plots display linear regression predictions of cognitive scores based on age and continuous plasma phosphorylated tau217 (p-tau217) represented with Ashton et al.^17^ three-variable categories. Predicted A) immediate and B) delayed recall scores were significantly by age by not by p-tau217. Age and p-tau217 both had a significant effect on C) semantic fluency (Supplementary Tables 4 5)

### Plasma p-tau217 by number of impaired cognitive scores

Next, we used delayed and semantic fluency measures to define cognitive status by the number of impaired cognitive tests to further investigate the association between ADNPC and cognitive status. The prevalence of ADNPC together with impairment in both delayed recall and semantic fluency was 10 and 13 % according to Figdore et al. and Ashton et al. dichotomic cut-offs, respectively. Prevalence of ADNPC was lowest (25.1–31.5 %) in participants with normative performance in both delayed recall and semantic fluency and highest (37.2–48.6 %) in participants with impaired performance in both tests (Figure 2).

**Figure 2:**
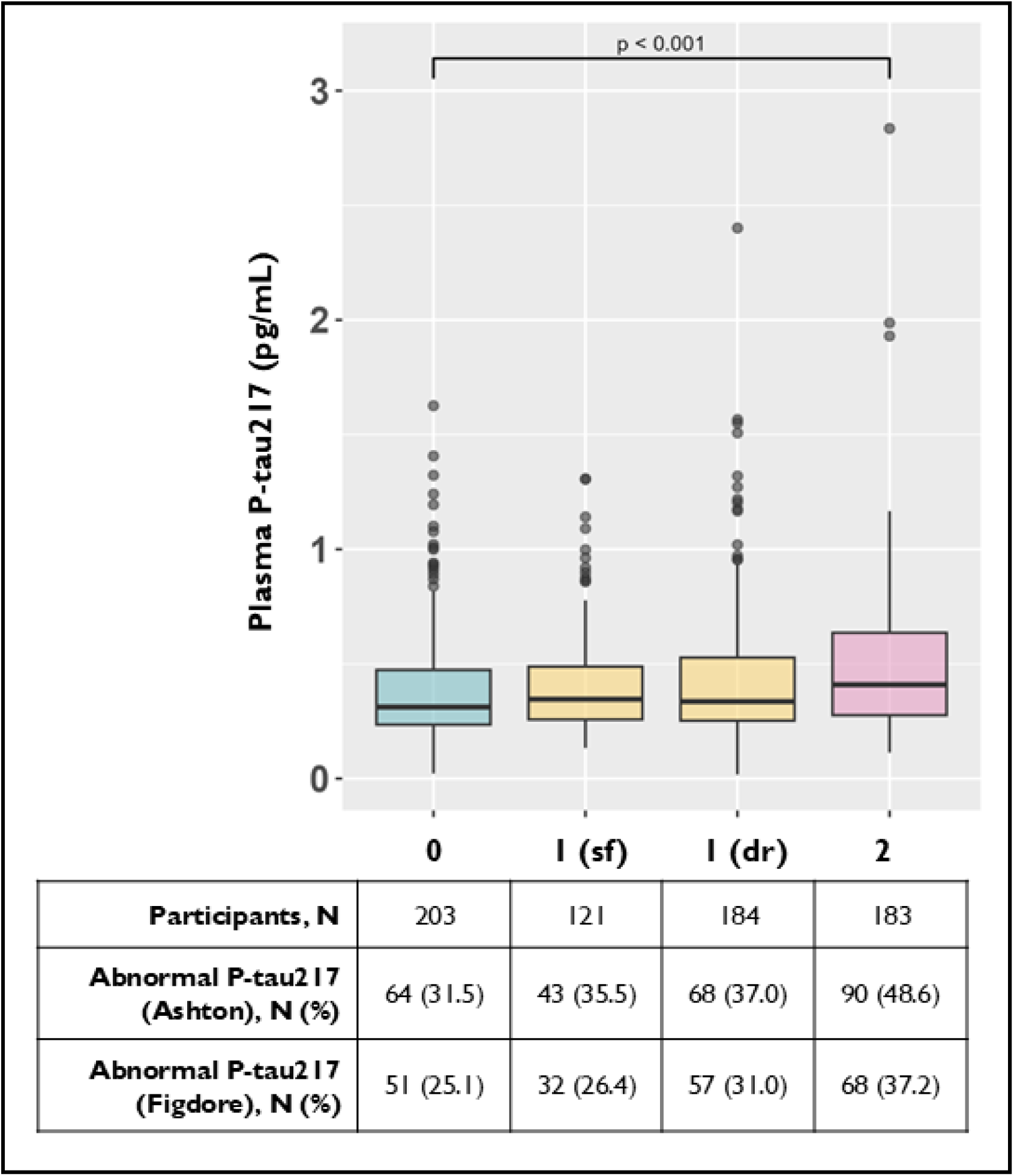
Prevalence of ADNPC by number of impaired delayed recall and semantic fluency cognitive scores. Plasma phosphorylated tau217 (p-tau217) status was analysed in cognitive groups defined with concurrent impairment in either or both semantic fluency (sf) and delayed recall (dr). Chi-squared test implied significant difference in Ashton et al. plasma p-tau217 abnormal level prevalence between the four groups, but not with Figdore et al. dichotomic categories (Supplementary Table 6). Continuous plasma p-tau217 levels differed significantly across the groups based on Wald-test, while significant difference in post-hoc tests was only found between groups with zero and two impaired scores (Supplementary Table 7).

The differences in ADNPC prevalence by dichotomic p-tau217 were significantly different between cognitive status groups when using Ashton^15^ (χ^2^ = 4.15, p = 0.006) but not using Figdore^17^ (χ^2^ = 2.55, p = 0.06) cut-offs (Supplementary Table 6). No differences were found when contrasting these groups against those who had impairment in only one test (Figure 2). Continuous plasma p-tau217 measures also differed significantly across cognitive status groups in Wald-test (W = 3.91, p = 0.009), and based on post hoc tests, we found a significant difference between groups with normal cognition and two impaired tests (t = 3.39, p < 0.001), while no significant differences were seen when comparing against those with only one impaired test (Figure 2, Supplementary Table 7).

## Discussion

This study set out to examine the prevalence of ADNPC and cognitive impairment using low-cost, scalable methods in a population-based sample without previously diagnosed dementia, and to examine the associations between cognitive performance and plasma p-tau217. Telephone tests measured episodic memory and semantic fluency, cognitive domains that are considered as the best predictors of progression to mild cognitive impairment and AD.^21^

We found 10—13 % of individuals without a clinical diagnosis of AD or other dementia-causing neurodegenerative disease exhibited concurrent episodic or semantic fluency impairment with amyloid positivity as measured with remote cognitive assessment and blood-samples, respectively. Individuals with impaired performance in delayed recall and semantic fluency had higher plasma p-tau217 levels than those who performed normally these AD sensitive tests. This supports the clinical approach of requiring impairment on more than one test, as an isolated abnormal score is common in neuropsychologically healthy individuals.^22^ Notably, a recent population-based study reported 34 % Aβ-positivity based on CSF Aβ42/40 ratio when median age was 63 years without cross-sectional p-tau217 association to cognition^23^, while we observed 30–38 % prevalence in an older sample with mean age 76 years depending on plasma p-tau217 cut-off with association to cognitive performance.

Despite the reported associations of p-tau217 and cognition, we note that a discrepancy in AD-related biomarker and cognitive status was common with approximately 40 % of participants had either ADNPC or impaired cognition. This is in line with earlier findings using PET and CSF measures of Aβ in those without dementia.^24^ In our study, the associations between continuous p-tau217 and cognitive measures were weak, highlighting the independence of ADNPC and cognition in individuals without a diagnosis of AD. The weak associations align with recent studies which suggest that plasma p-tau217 alone or with *APOE* and demographic data has low accuracy for detecting cognitive impairment.^6,25^ Both p-tau217 and episodic memory are age dependent, and here, the effect of age overshadowed the effect of p-tau217 on word list learning task measures. We observed great heterogeneity in plasma p-tau217 levels age-dependency in our sample of individuals without AD diagnosis.

Our results from population-based data are important for the ongoing discussion about the diagnosis of AD as a clinical-biological construct. Alzheimer’s association’s guidelines included plasma p-tau217 as one of the core biomarkers where abnormality is sufficient for diagnosing AD^4^, while determining AD in cognitively normal individuals based on biomarkers is not recommended^8^. However, the definition of cognitively normal in a real-world context is problematic due to the substantial prevalence of undiagnosed AD. In the clinical setting, cognitive normality is defined relative to age, sex and education. Yet, cognitive challenges are often the reason not to seek care, leading to detrimental down-stream effects for patients and clinical trials, as treatments benefit most those with the early observations of AD.^26^

Our findings indicate that combining plasma p-tau217 with telephone-administered cognitive assessment in individuals without a clinical AD diagnosis can help in classifying individuals in the AD continuum. Importantly, this approach was able to identify individuals who have both biologically defined AD and impairment in AD sensitive cognitive domains; this finding from a real-world sample of “cognitively normal” individuals suggest that using plasma p-tau217 and remote cognitive testing could improve the early detection of AD without imposing additional load on the healthcare system and would be widely accessible even without access to computers or smartphone apps.

### Strengths and limitations

Our study had several strengths. We used a population-based sample of older Finnish adults without a prior diagnosis of neurodegenerative disease, and we simultaneously analysed specific yet feasible ADNPC biomarkers and telephone-administered cognitive measures for early Alzheimer’s detection. Strength in the cognitive measures was that the episodic memory measures were validated in the TWINGEN sample and cut-offs were determined against in-person word list measures.^27^ Similarly, animal fluency cut-off was based on corresponding in-person administered test, but the validation has not been carried in the TWINGEN sample. Other studies have indicated validity of conducting this semantic fluency measure via telephone.^18,28^ A limitation is that cognitive cut-offs were based on the Finnish education-adjusted norms for 60-81-year-olds^19^in comparison to age range from 65 to 85 in our sample.

Finally, plasma p-tau217 literature is rapidly growing and several cut-offs for the ALZpath assay have been developed. With the lack of Finnish cut-offs, we used cut-offs based on two US samples^15,17^ the latter of which had a highly similar ethnicity to our sample. Alternative cut-offs are available and, for example, Schindler et al.^6^ reported 0.46 pg/mL cut-off for amyloid positivity that falls in between the cut-offs used in our study.

### Conclusions

A substantial minority of older adults without diagnosed dementia had a cognitive-biological profile consistent with AD in this population-based sample. Our results indicate that detecting individuals with undiagnosed or high risk for AD is possible by combining the scalable, low-cost methods of plasma p-tau217 and remote cognitive assessment via a short telephone interview.

## Data Availability

In accordance with the Finnish Biobank Act, the data used in the analysis is deposited in the Biobank of the Finnish Institute for Health and Welfare. (https://thl.fi/en/web/thl-biobank/forresearchers). It is available to researchers from academia and companies after a written application and following the relevant Finnish legislation. To ensure the protection of privacy and compliance with national data protection legislation, a data use/transfer agreement is needed, the content and specific clauses of which will depend on the nature of the requested data.

## Acknowledgements

We thank the participants of the older Finnish Twin Cohort study who participated in NONAGINTA and TWINGEN studies. The participants of the TWINGEN study were recruited through THL Biobank (study number THLBB2022_83). We thank all participants for their participation in biobank research. We thank Auli Toivola and Aija Kyttälä from the THL biobank for study coordination and participant recall in TWINGEN. We also thank Helsinki Biobank, Finnish Clinical Biobank Tampere, Arctic Biobank, Biobank Borealis of Northern Finland, Central Finland Biobank, Biobank of Eastern Finland and Turku University of Applied Sciences for their contributions to TWINGEN data collection. The biobank personnel who carried out the data collection and processing of the samples are the following: Sabrina Belgasem (Helsinki Biobank), Henna Palin, Minttu Virolainen, and Anna-Kaisa Pohjonen (Finnish Clinical Biobank Tampere), Anu Outinen-Tuuponen, Marja-Leena Kytökangas, and Riikka-Mari Siiro-Virtanen (Arctic Biobank and Biobank Borealis of Northern Finland), Senni Lipponen (Central Finland Biobank), Nina Hurula (Biobank of Eastern Finland), and biomedical laboratory scientist students Heidi Kalve and Anniina Friman (Turku University of Applied Sciences’ clinical laboratory). We thank Tarja Hallaranta (University of Helsinki) for conducting telephone interviews for data collection. We thank Jyrki Tammerluoto, Steffi Besselink, Huei-Yi Shen, and Anu Jalanko for administrative work.

---

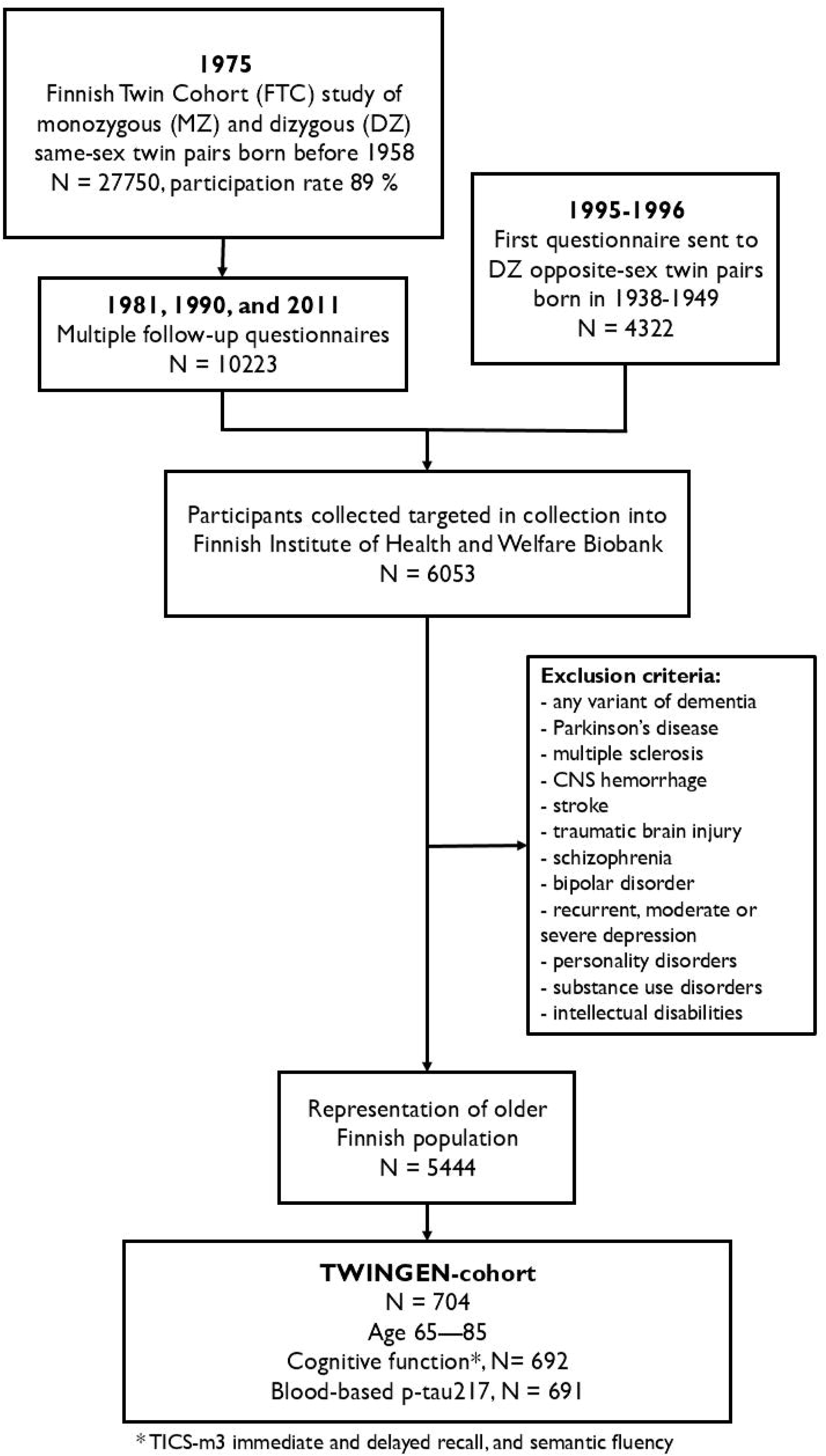

