## Supplementary Material for "Scalable biological-cognitive profiling for Alzheimer’s disease in the population"

**Scalable biological-cognitive profiling for Alzheimer’s disease in the population**
S**upplementary Figure 1. Flowchart of original twin participant recruitment and selection to TWINGEN-cohort.^[1]^**

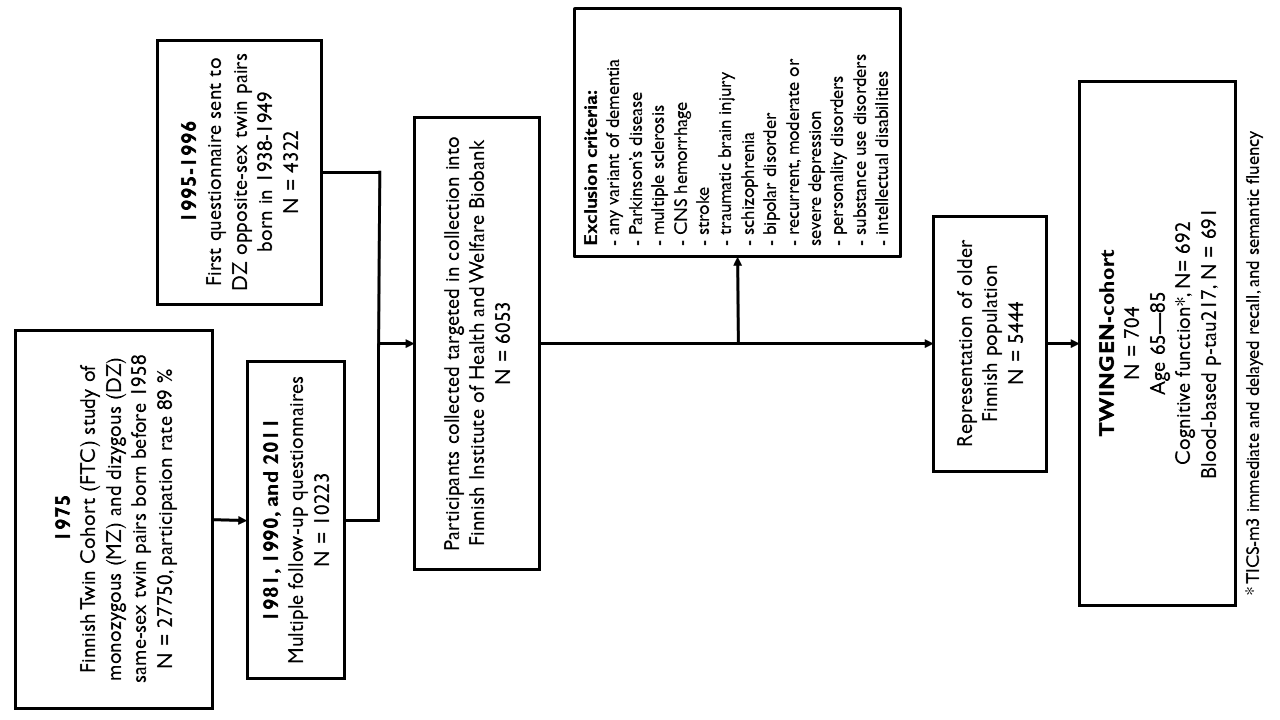

**Supplementary Table 1: Cognitive performance variance by Figdore et al. p-tau217 categories.**

|  | | **Dichotomic Plasma P-tau217** | | | **Three-category Plasma P-tau217** | | | |
| --- | --- | --- | --- | --- | --- | --- | --- | --- |
|  | | **Normal** | **Abnormal** | **Statistical Difference**  (statistic [df^[2]^/ddf^[3]^], p-value) | **Low** | **Intermediate** | **High** | **Statistical Difference** (statistic [ddf], p-value) |
|  | **N** | 483 | 208 |  | 411 | 123 | 157 |  |
|  | **P-tau217 pg/ml, Median (IQR)** | 0.29 (0.13) | 0.73 (0.32) |  | 0.27 (0.11) | 0.46 (0.07) | 0.82 (0.30) |  |
| **Immediate**  **Recall** | **Score, M (SD)** | 17.66 (4.21) | 16.85 (4.22) | t = 2.25 (df = 538), p = 0.03 | 17.71 (4.19) | 17.01 (4.28) | 16.97 (4.26) | W = 2.20 (537), p = 0.11 |
|  | **Impaired Cognition, N (%)** | 211 (43.7) | 112 (53.8) | χ^2^ = 5.66 (ddf = 539), p = 0.02 | 174 (42.3) | 64 (52.0) | 85 (54.1) | χ^2^ = 3.82 (1077), p = 0.02 |
|  | **Score > AD score, N (%)** | 326 (74.9) | 138 (66.3) | χ^2^ = 5,28 (ddf = 539), p = 0.02 | 311 (75.7) | 85 (69.1) | 104 (66.2) | χ^2^ = 2.82 (1078), p = 0.06 |
| **Delayed**  **Recall** | **Score, M (SD)** | 4.92 (2.22) | 4.40 (2.46) | t = 2.50 (df = 538), p = 0.01 | 5.00 (2.20) | 4.50 (2.24) | 4.34 (2.54) | W = 4.63 (537), p = 0.01 |
|  | **Impaired Cognition, N (%)** | 242 (50.1) | 125 (60.1) | χ^2^ = 5.50 (ddf = 539), p = 0.02 | 202 (49.1) | 69 (56.1) | 96 (61.1) | χ^2^ = 3.47 (1072), p = 0.03 |
|  | **Score > AD score, N (%)** | 347 (71.8) | 138 (66.3) | χ^2^ = 2.03 (ddf = 539), p = 0.15 | 302 (73.5) | 82 (66.7) | 101 (64.3) | χ^2^ = 2.68 (1077), p = 0.07 |
| **Semantic**  **Fluency** | **Score, M (SD)** | 19.18 (5.28) | 17.91 (4.79) | t = 2.96 (df = 538), p = 0.003 | 19.51 (5.12) | 17.99 (5.51) | 17.59 (4.71) | W = 9.53 (537), p < 0.001 |
|  | **Impaired Cognition, N (%)** | 204 (42.2) | 100 (48.1) | χ^2^ = 1.92 (ddf = 539), p = 0.17 | 163 (39.7) | 64 (52.0) | 77 (49.0) | χ^2^ = 3.82 (1077), p = 0.02 |
|  | **Score > AD score, N (%)** | 358 (74.1) | 144 (69.2) | χ^2^ = 4.50 (ddf = 539), p = 0.03 | 317 (77.1) | 78 (63.4) | 107 (68.2) | χ^2^ = 2.97 (1078), p = 0.05 |

|  | | | **Three-category plasma p-tau217 and Cognitive Status** (statistic [DF], adjusted p-value^[4]^) | | |
| --- | --- | --- | --- | --- | --- |
|  |  |  | **Low-Intermediate** | **Low-High** | **Intermediate-High** |
| **Ashton et al.** | **Immediate Recall** | **Score** | t = -1.56 (441), p = 0.18 | t = -1.92 (444), p = 0.06 | t = -0.36 (248), p = 0.72 |
|  |  | **Impaired Cognition** | χ^2^ = 3.44 (442), p = 0.18 | χ^2^ = 6.32 (445), p = 0.03 | χ^2^ = 0.38 (249), p = 0.72 |
|  |  | **Score > AD score** | χ^2^ = 1.81 (442), p = 0.18 | χ^2^ = 5.58 (445), p = 0.03 | χ^2^ = 0.67 (249), p = 0.72 |
|  | **Delayed**  **Recall** | **Score** | t = -2.07 (441), p = 0.12 | t = -2.73 (444), p = 0.02 | t = -0.86 (248), p = 0.49 |
|  |  | **Impaired Cognition** | χ^2^ = 2.61 (442), p = 0.16 | χ^2^ = 5.16 (445), p = 0.02 | χ^2^ = 0.47 (249), p = 0.49 |
|  |  | **Score > AD score** | χ^2^ = 1.70 (442), p = 0.19 | χ^2^ = 5.15 (445), p = 0.02 | χ^2^ = 0.72 (249), p = 0.49 |
|  | **Semantic Fluency** | **Score** | t = -3.03 (441), p = 0.01 | t = -3.98 (444), p < 0.001 | t = -0.44 (248), p = 0.71 |
|  |  | **Impaired Cognition** | χ^2^ = 7.79 (442), p = 0.01 | χ^2^ = 2.30 (445), p = 0.13 | χ^2^ = 1.09 (249), p = 0.71 |
|  |  | **Score > AD score** | χ^2^ = 2.94 (442), p = 0.09 | χ^2^ = 4.42 (445), p = 0.05 | χ^2^ = 0.14 (249), p = 0.71 |
| **Figdore et al.** | **Immediate Recall** | **Score** | t = -1.57 (427), p = 0.16 | t = -1.77 (463), p = 0.08 | t = -0.08 (245), p = 0.94 |
|  |  | **Impaired Cognition** | χ^2^ = 3.42 (428), p = 0.16 | χ^2^ = 5.95 (464), p = 0.04 | χ^2^ = 0.12 (246), p = 0.94 |
|  |  | **Score > AD score** | χ^2^ = 2.01 (428), p = 0.16 | χ^2^ = 5.05 (464), p = 0.04 | χ^2^ = 0.25 (246), p = 0.94 |
|  | **Delayed**  **Recall** | **Score** | t = -2.08 (427), p = 0.11 | t = -2.68 (463), p = 0.02 | t = -0.55 (245), p = 0.68 |
|  |  | **Impaired Cognition** | χ^2^ = 1.89 (428), p = 0.17 | χ^2^ = 5.99 (464), p = 0.02 | χ^2^ = 0.74 (246), p = 0.68 |
|  |  | **Score > AD score** | χ^2^ = 2.18 (428), p = 0.17 | χ^2^ = 4.40 (464), p = 0.04 | χ^2^ = 0.17 (246), p = 0.68 |
|  | **Semantic Fluency** | **Score** | t = -2.68 (427), p = 0.02 | t = -4.06 (463), p < 0.001 | t = -0.64 (245), p = 0.88 |
|  |  | **Impaired Cognition** | χ^2^ = 5.67 (428), p = 0.03 | χ^2^ = 3.86 (464), p = 0.05 | χ^2^ = 0.24 (246), p = 0.88 |
|  |  | **Score > AD score** | χ^2^ = 3.12 (428), p = 0.08 | χ^2^ = 4.46 (464), p = 0.05 | χ^2^ = 0.02 (246), p = 0.88 |

**Supplementary Table 2: Post-Hoc Test for cognitive scores, prevalence of cognitive impairment, and prevalence of impairment corresponding to average AD patient’s performance by three-category plasma p-tau217 groups.**

**Supplementary Table 3: Spearman-correlations of age, education, and plasma p-tau217 with cognitive scores**.

|  | | **Pearson Correlations** | |
| --- | --- | --- | --- |
|  |  | **Age** | **Plasma P-tau217** |
| **Immediate** | **Coefficient (SE)** | r = - 0.127 (0.039) | r = - 0.083 (0.040) |
|  | **Statistics** | t = - 3.27 (p < 0.001) | t = - 2.03 (p = 0.052) |
| **Delayed** | **Coefficient (SE)** | r = - 0.202 (0.037) | r = - 0.113 (0.039) |
|  | **Statistics** | t = - 5.52 (p < 0.001) | t = - 2.86 (p < 0.006) |
| **Semantic Fluency** | **Coefficient (SE)** | r = - 0.149 (0.040) | r = - 0.167 (0.032) |
|  | **Statistics** | t = - 3.71 (p < 0.001) | t = - 5.23 (p < 0.001) |

**Supplementary Table 4: Linear regression predictions for cognition with age, p-tau217 as covariates.**

|  |  | **Estimate** | **95 % CI** | **SE** | **T-value** | **Adjusted P-value** |
| --- | --- | --- | --- | --- | --- | --- |
| **Immediate Recall** | **(Intercept)** | 17.056 | 16.348, 17.764 | 0.360 | 47.325 | < 0.001 |
|  | **Age** | -0.476 | -0.871, -0.080 | 0.201 | -2.362 | 0.028 |
|  | **P-tau217** | -0.386 | -1.055, 0.282 | 0.340 | -1.135 | 0.257 |
| **Delayed Recall** | **(Intercept)** | 4.523 | 4.142, 4.904 | 0.194 | 23.312 | < 0.001 |
|  | **Age** | -0.427 | -0.618, -0.235 | 0.097 | -4.385 | < 0.001 |
|  | **P-tau217** | -0.253 | -0.596, 0.089 | 0.174 | -1.451 | 0.147 |
| **Semantic fluency** | **(Intercept)** | 17.631 | 16.905, 18.358 | 0.370 | 47.675 | < 0.001 |
|  | **Age** | -0.576 | -1.035, -0.117 | 0.234 | -2.464 | 0.014 |
|  | **P-tau217** | -1.251 | -1.916, -0.587 | 0.338 | -3.701 | < 0.001 |

**Supplementary Table 5: Linear regression predictions for cognition with age, p-tau217 and age x p-tau217 interaction as covariates.**

|  |  | **Estimate** | **95 % CI** | **SE** | **T-value** | **Adjusted P-value** |
| --- | --- | --- | --- | --- | --- | --- |
| **Immediate Recall** | **(Intercept)** | 17.001 | 16.310, 17.692 | 0.352 | 48.317 | < 0.001 |
|  | **Age** | -0.148 | -0.829, 0.533 | 0.347 | -0.427 | 0.670 |
|  | **P-tau217** | -0.391 | -1.041, 0.258 | 0.331 | -1.183 | 0.383 |
|  | **Age:P-tau217** | 0.319 | -0.270, 0.908 | 0.300 | 1.065 | 0.383 |
| **Delayed Recall** | **(Intercept)** | 4.515 | 4.141, 4.889 | 0.190 | 23.729 | < 0.001 |
|  | **Age** | -0.377 | -0.743, -0.012 | 0.186 | -2.027 | 0.086 |
|  | **P-tau217** | -0.254 | -0.594, 0.086 | 0.173 | -1.468 | 0.190 |
|  | **Age:P-tau217** | 0.048 | -0.259, 0.355 | 0.156 | 0.307 | 0.759 |
| **Semantic fluency** | **(Intercept)** | 17.530 | 16.783, 18.276 | 0.380 | 46.122 | < 0.001 |
|  | **Age** | 0.031 | -0.744, 0.805 | 0.394 | 0.078 | 0.938 |
|  | **P-tau217** | -1.261 | -1.927, -0.596 | 0.339 | -3.724 | < 0.001 |
|  | **Age:P-tau217** | 0.591 | -0.036, 1.217 | 0.319 | 1.853 | 0.086 |

**Supplementary Table 6: Number of impaired scores based on delayed recall and semantic fluency.**^[5]^

|  |  | | **Number of Impaired Scores** | | | | **Statistical Difference** | |
| --- | --- | --- | --- | --- | --- | --- | --- | --- |
|  |  |  | **0** | **1 (sf)**^[6]^ | **1 (dr)**^[7]^ | **2** | **χ^2^-Statistic (DF)** | **P-value** |
| **Ashton et al.** | **Dichotomic plasma p-tau217** | **Normal** (n = 427) | 139 | 78 | 116 | 94 | 4.15 (1611) | 0.006 |
|  |  | **Abnormal** (n = 265) | 64 | 43 | 68 | 89 |  |  |
|  | **Three-category plasma p-tau217** | **Low** (n = 407) | 135 | 72 | 111 | 89 | 2.55 (3204) | 0.02 |
|  |  | **Intermediate** (n = 146) | 33 | 30 | 35 | 48 |  |  |
|  |  | **High** (n = 138) | 35 | 19 | 38 | 46 |  |  |
| **Figdore et al.** | **Dichotomic**  **plasma p-tau217** | **Normal** (n = 483) | 152 | 89 | 127 | 115 | 2.47 (1613) | 0.06 |
|  |  | **Abnormal** (n = 208) | 51 | 32 | 57 | 68 |  |  |
|  | **Three-category plasma p-tau217** | **Low** (n = 411) | 135 | 74 | 113 | 89 | 2.44 (3204) | 0.02 |
|  |  | **Intermediate** (n = 123) | 31 | 23 | 28 | 41 |  |  |
|  |  | **High** (n = 157) | 37 | 24 | 43 | 53 |  |  |

**Supplementary table 7: Post-hoc Two-tailed T-tests of Plasma P-tau217 levels by cognitive performance in Delayed Recall and Semantic Fluency**

|  | **Two-tailed T-test on Plasma P-tau217 level** | |
| --- | --- | --- |
|  | **T-statistic (DF)** | **Adjusted P-value**^[8]^ |
| **0 – 1 (sf)** | 1.62 (276) | 0.10 |
| **0 – 1 (dr)** | 1.20 (319) | 0.23 |
| **0 – 2** | 3.39 (328) | < 0.001 |
| **1 (sf) – 1 (dr)** | -0.29 (274) | 0.77 |
| **1 (sf) – 2** | 1.77 (266) | 0.08 |
| **1 (dr) – 2** | 1.99 (315) | 0.05 |

1. Finnish Twin Cohort study (FTC) was initiated in 1975, when invitation for the study was sent to all same-sex twins born before 1958. These same-sex twin pairs were followed with multiple questionnaires in 1981 and 2017. Opposite-sex twin pairs born during same decades were contacted later and received baseline questionnaire between 1995 and 1996. Later, participants who participated to FTC study were contacted and their information was collected into Finnish Institute of Health and Welfare Biobank across multiple biobanks located in Finland after excluding participants with clinically diagnosed neurodegenerative disease or other cognition- or personality-affecting traumatic or chronic diseases. TWINGEN-cohort is a carefully selected representative population of these individuals, that reflects age related cognitive changes as the participants underwent multiple cognitive assessments in-clinical and on telephone and were analysed for plasma p-tau217.
2. Degrees of freedom
3. Denominator degrees of freedom
4. Benjamini-Hochberg procedure applied on all p-values
5. Participant groups based on number of impaired cognitive tests of semantic fluency (sf) and delayed recall (dr).
6. Semantic fluency
7. Delayed recall
8. Benjamini-Hochberg procedure applied on all p-values
